# Parental plans to vaccinate children for COVID-19 in New York City

**DOI:** 10.1101/2021.05.26.21257871

**Authors:** Chloe A. Teasdale, Luisa N. Borrell, Yanhan Shen, Spencer Kimball, Michael L. Rinke, Sasha A. Fleary, Denis Nash

## Abstract

Once COVID-19 vaccines are approved for children <12 years of age, high pediatric vaccination coverage will be needed to help minimize the public health threat from the SARS-CoV-2 epidemic. We conducted an online survey of 1,119 parents and caregivers of children ≤12 years in New York City from March 9 to April 11, 2021. Among parents surveyed, 61.9% reported plans to vaccinate their youngest child for COVID-19, 14.8% said they do not plan to vaccinate their child and 23.3% were unsure. Female and non-Hispanic Black parents were least likely to report plans to vaccinate their children. Safety, effectiveness and perceptions that children do not need vaccination were the primary reasons for vaccine hesitancy/resistance. Parents who have or will vaccinate themselves were significantly more likely to report they would vaccinate their children. Efforts to increase awareness about vaccine safety and education about the importance of vaccinating children are needed.

---

As of May 2021, vaccines to prevent SARs-COV-2 infection (COVID-19) have not yet been approved for use in children younger than 12 years of age in the United States (US). Clinical trials in younger pediatric populations are underway and approval for vaccines in children under 12 years is expected before the end of 2021.^1^ Once COVID-19 vaccination is available for children, acceptability among parents will be critical for protecting them from infection and for mitigating the public health threat posed by COVID-19.^2^

While children may be a lower risk for severe disease, almost 4 million children in the US have been diagnosed with COVID-19 since the start of the epidemic and approximately 300 have died.^3^ In addition, some infected children have serious complications, including multi□inflammatory syndrome in children (MIS□C), and children may also experience long COVID symptoms similar to adults.^4,5^

Preventing COVID-19 in children will not only have individual health benefits, it will also contribute to stopping the spread of infections in communities and households. Although children appear to be less susceptible to infection and to play a smaller role than teenagers and adults in transmission,^2^ there have been outbreaks reported from schools and daycare facilities^6,7^ and age-appropriate social distancing measures are recommended for children.^8^ High coverage of COVID-19 vaccination among children will help stop transmission and will allow for safe return to normal activities, including fully opening schools.^9^

New York City (NYC) was the epicenter of the US COVID-19 epidemic and continues to have high levels of community transmission.^10^ It is the most densely populated city in the US and also one of the most racially and ethnicity diverse. Since the start of the epidemic, more than 87,000 children in NYC have been diagnosed with COVID-19^3^ and, in an analysis of NYC testing data, school aged children 5-14 years who were tested for SARS-CoV-2 had the highest sero-prevalence of any age group.^11^

## METHODS

We conducted an online non-probability survey of English and Spanish speaking parents and caregivers (‘parents’) in NYC to measure intentions to vaccinate their youngest child when a pediatric COVID-19 vaccine is available (n=1,119). Eligible participants were adults ≥18 years who identified as primary caregivers of a child ≤12 years of age. Recruitment was conducted through a Qualtrics panel with data collected from March 9 through April 11, 2021. We followed the American Association for Public Opinion Research (AAPOR) guidelines for quota-based sampling^12^ and used 2019 Census data for NYC to calculate survey weights based on race, ethnicity, sex, education and borough.^13^ The protocol was approved by the CUNY School of Public Health and Health Policy institutional review board.

The study outcome was the proportion of parents reporting intentions to vaccinate the youngest child in their household against COVID-19. Parents were asked “when a vaccine to prevent COVID-19 is approved for children, would you want your child to receive the vaccine” (responses: “yes”, “no”, “unsure”). Parents responding “no” or “unsure” were considered vaccine hesitant/resistant and were asked, “why do you not want your child to receive the COVID-19 vaccine?” with multiple response options. Parents reported demographics and household characteristics, and whether they had received or planned to get the COVID-19 vaccine themselves (response options included already received COVID-19 vaccination, plan to receive when available, unsure, will not get the vaccine and prefer not to answer).

Survey weights were applied to all analyses to generate prevalence estimates. Descriptive statistics (unweighted counts and weighted percentages) are presented for the sample. Prevalence of parental plans to vaccinate children (yes, no, unsure) were compared by sample characteristics using Rao adjusted Pearson chi-squared tests. Poisson regression models with robust standard errors were fitted to estimate prevalence ratios (PR) and confidence intervals (CI) comparing parents planning to vaccinate to those responding “no” or “unsure” (combined); models were adjusted for demographic and household characteristics. Parents’ own COVID-19 vaccination status was examined by grouping together parents who responded that they had already been vaccinated with those who planned to get the vaccine, and those responding that they were unsure or preferred not to answer (parents reporting they would not get vaccinated were examined as their own group). We measured the association between parental vaccine status with reported intentions to vaccinate children using Rao adjusted Pearson chi-squared tests to compare proportions. Analyses were conducted in SAS 9.4 (SAS Institute Inc., Cary, NC, USA).

## RESULTS

Among NYC parents, 61.9% reported intentions to vaccinate their youngest child against COVID-19 (median child age: 4.7 years), 14.8% said they will not vaccinate their child and 23.3% were unsure (Table 1). The most commonly cited concerns among vaccine hesitant parents (those reporting they would not vaccinate their child or were unsure) were safety and effectiveness which was reported by 81.2%, and there were significant differences by race/ethnicity. Whereas 88.4% of Hispanic parents, 85.1% of non-Hispanic Black parents and 82.3% of Asian parents who were vaccine hesitant reported safety and effectiveness concerns, only 60.7% of non-Hispanic white vaccine-hesitant parents reported this (p=0.01). In addition to safety concerns, 21.7% of vaccine hesitant parents reported that children are at low risk for COVID-19 and don’t need vaccination; 16.6% and 9.5% of vaccine hesitant parents reported medical and religious or philosophical reasons, respectively, and 2.7% reported ‘other’ reasons.

**Table 1.** Intentions among NYC parents to vaccinate children ≤12 years against COVID-19 according to parent and child characteristics –. March 9-April 8, 2021

|  |  |  | When a COVID-19 vaccine available for children, will you want your child to be vaccinated |  |  |  |  |  |  |  |  |  |
| --- | --- | --- | --- | --- | --- | --- | --- | --- | --- | --- | --- | --- |
|  | NYC sample |  | Yes |  | No |  | Unsure |  | p-value† | Adjusted prevalence ratios |  |  |
| Characteristics | N | %* | %^ | 95% CI | % | 95% CI | % | 95% CI |  | aPR | 95%CI | p-value |
| Total sample | 1119 | 100.0 | 61.9 | 56.6-67.1 | 14.8 | 11.1-18.6 | 23.3 | 18.4-28.3 |  |  |  |  |
| Child age |  |  |  |  |  |  |  |  |  |  |  |  |
| Median age, years (IQR) | 4.7 (2.0-8.5) |  | 4.9 (2.2-8.5) |  | 5.6 (2.1-8.6) |  | 3.5 (1.4-8.2) |  | <0.01 |  |  |  |
| <24 months | 195 | 18.4 | 55.4 | 41.0-69.7 | 14.8 | 4.4-25.3 | 29.8 | 14.6-45.0 | 0.59 | 1.00 | ref | -- |
| 2-6 years | 460 | 41.1 | 64.2 | 56.5-71.8 | 12.5 | 7.8-17.1 | 23.3 | 16.1-30.6 |  | 1.03 | 0.82-1.30 | 0.76 |
| 7-12 years | 464 | 40.5 | 62.4 | 54.8-70.1 | 17.2 | 10.9-23.5 | 20.4 | 13.8-27.0 |  | 1.03 | 0.81-1.30 | 0.81 |
| Child sex |  |  |  |  |  |  |  |  |  |  |  |  |
| Male | 558 | 50.8 | 62.8 | 54.8-70.9 | 12.4 | 6.6-18.1 | 24.8 | 17.2-32.4 | 0.09 | 1.00 | ref | -- |
| Female | 555 | 48.7 | 61.6 | 54.7-68.4 | 17.1 | 12.2-22.0 | 21.4 | 14.9-27.9 |  | 1.01 | 0.88-1.16 | 0.85 |
| Missing§ | 6 | 0.5 |  |  |  |  |  |  |  |  |  |  |
| Child race/ethnicity** |  |  |  |  |  |  |  |  |  |  |  |  |
| Black non-Hispanic | 162 | 18.4 | 45.8 | 36.2-55.4 | 23.2 | 15.7-30.8 | 31.0 | 22.3-39.7 | <0.01 |  |  |  |
| Asian | 100 | 12.8 | 67.9 | 55.8-80.0 | 5.0 | 0.9-9.1 | 27.1 | 15.2-39.0 |  |  |  |  |
| White non-Hispanic | 517 | 31.7 | 86.6 | 82.5-90.8 | 7.2 | 4.4-10.1 | 6.1 | 2.8-9.4 |  |  |  |  |
| Hispanic | 294 | 29.3 | 44.8 | 32.1-57.4 | 19.6 | 9.5-29.8 | 35.6 | 22.5-48.8 |  |  |  |  |
| Other non-Hispanic§ | 46 | 7.6 |  |  |  |  |  |  |  |  |  |  |
| Parent age |  |  |  |  |  |  |  |  |  |  |  |  |
| 18-29 years | 165 | 23.2 | 40.9 | 28.0-53.9 | 19.0 | 7.5-30.5 | 40.1 | 25.5-54.6 | <0.01 | 0.87 | 0.61-1.24 | 0.44 |
| 30-44 years | 830 | 67.4 | 69.1 | 63.9-74.2 | 12.7 | 9.2-16.3 | 18.2 | 13.7-22.7 |  | 1.01 | 0.81-1.26 | 0.93 |
| 45+ years | 124 | 9.4 | 61.7 | 48.7-74.6 | 19.4 | 8.5-30.3 | 19.0 | 9.2-28.7 |  | 1.00 | ref | -- |
| Parent sex |  |  |  |  |  |  |  |  |  |  |  |  |
| Male | 626 | 40.7 | 83.3 | 78.3-88.2 | 6.6 | 3.8-9.5 | 10.1 | 5.8-14.5 | <0.01 | 1.00 | ref | -- |
| Female | 490 | 59.0 | 47.2 | 39.8-54.7 | 20.4 | 14.4-26.3 | 32.4 | 24.9-39.9 |  | 0.72 | 0.61-0.85 | <0.0001 |
| Transgender/other§# | 3 | 0.3 |  |  |  |  |  |  |  |  |  |  |
| Parent race/ethnicity |  |  |  |  |  |  |  |  |  |  |  |  |
| Black non-Hispanic | 176 | 20.9 | 47.3 | 38.2-56.4 | 23.4 | 16.1-30.8 | 29.3 | 21.2-37.3 | <0.01 | 0.79 | 0.63-0.99 | 0.04 |
| Asian | 113 | 14.2 | 67.4 | 56.1-78.8 | 5.0 | 1.2-8.9 | 27.5 | 16.3-38.7 |  | 1.05 | 0.84-1.31 | 0.67 |
| White non-Hispanic | 510 | 31.1 | 86.2 | 82.0-90.4 | 7.6 | 4.7-10.5 | 6.2 | 2.9-9.6 |  | 1.00 | ref | -- |
| Hispanic | 292 | 29.2 | 47.0 | 33.8-60.2 | 19.2 | 9.0-29.4 | 33.8 | 20.4-47.2 |  | 0.80 | 0.61-1.05 | 0.11 |
| Other non-Hispanic | 28 | 4.7 | 40.7 | 16.8-64.6 | 26.7 | 3.5-50.0 | 32.6 | 11.2-53.9 |  | 0.72 | 0.41-1.26 | 0.24 |
| Child has health insurance |  |  |  |  |  |  |  |  |  |  |  |  |
| Yes | 1018 | 91.3 | 61.0 | 55.5-66.6 | 15.2 | 11.2-19.2 | 23.8 | 18.5-29.1 | 0.14 | 0.99 | 0.81-1.20 | 0.91 |
| No | 89 | 6.3 | 78.5 | 67.3-89.7 | 11.6 | 2.8-20.5 | 9.9 | 2.0-17.7 |  | 1.00 | ref | -- |
| Don't know | 12 | 2.4 |  |  |  |  |  |  |  |  |  |  |
| **Child attending in person school/daycare ≥1 day per week** |  |  |  |  |  |  |  |  |  |  |  |  |
| Yes | 628 | 50.7 | 71.5 | 64.4-78.5 | 11.8 | 7.6-16.0 | 16.7 | 10.0-23.5 | <0.01 | 1.23 | 1.05-1.45 | 0.01 |
| No | 482 | 48.3 | 52.5 | 44.9-60.1 | 18.0 | 11.7-24.2 | 29.5 | 22.2-36.9 |  | 1.00 | ref | -- |
| Don't know§ | 9 | 1.0 |  |  |  |  |  |  |  |  |  |  |
| **Number of children ≤12 years in household** |  |  |  |  |  |  |  |  |  |  |  |  |
| 1 | 514 | 49.4 | 55.8 | 48.6-62.9 | 14.4 | 10.0-18.8 | 29.8 | 22.7-36.9 | 0.07 | 1.00 | ref | -- |
| 2 | 494 | 40.2 | 70.0 | 61.2-78.8 | 13.6 | 7.4-19.8 | 16.4 | 8.2-24.7 |  | 1.06 | 0.89-1.26 | 0.50 |
| 3 or more | 111 | 10.4 | 59.3 | 42.1-76.4 | 21.4 | 5.0-37.9 | 19.3 | 8.6-29.9 |  | 1.04 | 0.77-1.42 | 0.79 |
| **Parent education (highest completed)** |  |  |  |  |  |  |  |  |  |  |  |  |
| High school or less | 145 | 30.0 | 46.9 | 34.6-59.2 | 19.2 | 9.5-29.0 | 33.9 | 21.4-46.3 | <0.01 | 0.90 | 0.69-1.16 | 0.40 |
| Some college/tech school | 193 | 30.3 | 57.6 | 47.9-67.2 | 15.1 | 8.4-21.8 | 27.4 | 18.3-36.4 |  | 1.02 | 0.83-1.25 | 0.84 |
| Completed college or more | 761 | 38.4 | 76.1 | 72.2-80.1 | 11.4 | 8.5-14.3 | 12.5 | 9.4-15.6 |  | 1.00 | ref | -- |
| Missing§ | 20 | 1.4 |  |  |  |  |  |  |  |  |  |  |
| **Household income (USD)** |  |  |  |  |  |  |  |  |  |  |  |  |
| <\$25,000 | 106 | 15.1 | 52.0 | 36.8-67.2 | 16.7 | 8.7-24.6 | 31.3 | 16.8-45.9 | <0.01 | 0.74 | 0.53-1.03 | 0.08 |
| \$25,000-\$49,999 | 157 | 20.8 | 38.3 | 26.0-50.7 | 22.3 | 11.0-33.7 | 39.4 | 25.2-53.5 | | 0.98 | 0.86-1.13 | 0.82 |
| \$50,000-\$99,999 | 275 | 26.8 | 62.5 | 52.2-72.8 | 15.2 | 7.0-23.3 | 22.3 | 13.3-31.3 | | 1.13 | 0.80-1.60 | 0.48 |
| ≥\$100,000 | 554 | 31.7 | 82.1 | 77.3-86.8 | 8.9 | 4.8-13.1 | 9.0 | 6.0-12.1 | | 1.00 | ref | -- |
| Missing§ | 27 | 5.6 |  |  |  |  |  |  |  |  |  |  |
| **NYC borough** |  |  |  |  |  |  |  |  |  |  |  |  |
| Bronx | 278 | 13.7 | 48.3 | 38.6-57.9 | 21.9 | 14.3-29.4 | 29.9 | 20.4-39.3 | <0.01 | 0.92 | 0.79-1.06 | 0.26 |
| Brooklyn | 360 | 31.7 | 63.1 | 55.1-71.1 | 17.6 | 11.2-24.0 | 19.3 | 12.3-26.3 |  | 1.01 | 0.81-1.25 | 0.95 |
| Manhattan | 240 | 14.9 | 83.7 | 75.4-92.1 | 5.1 | 1.0-9.1 | 11.2 | 3.4-18.9 |  | 1.00 | ref | -- |
| Staten Island | 46 | 8.0 | 57.6 | 32.4-82.9 | 7.3 | 0.4-14.1 | 35.1 | 8.4-61.8 |  | 0.85 | 0.60-1.21 | 0.36 |
| Queens | 195 | 31.6 | 57.3 | 46.1-68.5 | 15.4 | 6.6-24.2 | 27.3 | 16.9-37.7 |  | 0.84 | 0.66-1.06 | 0.14 |

Abbreviations: CI = confidence interval; IQR = interquartile range; USD = US dollars.
\*Survey weights applied to sample to represent NYC population of parents by race, ethnicity, sex, education and region.
^Weighted percentages are prevalence estimates of NYC parents reporting vaccination plans for their youngest child.
‡p-values from Rao adjusted Pearson Chi-squared tests to compare expected to observed frequencies among groups by characteristic for parent's willingness to vaccinate their youngest child (i.e. whether willing to vaccinate youngest child differed by sex of the child, etc.);
§Categories are not presented in the table as they yielded unreliable standard error estimates.
\*\*Child's race/ethnicity excluded from adjusted models due to collinearity with parent's race/ethnicity.

In adjusted models, parents of children attending in-person school or daycare were more likely to report intentions to vaccinate (aPR: 1.23;95% confidence interval (CI): 1.05-1.45) (Table 1). Female (aPR: 0.72;95%CI: 0.61-0.85) and non-Hispanic Black parents (aPR: 0.79, 95%CI: 0.63-0.99) were less likely to report plans to vaccinate their children.

Among parents, 20.2% had received and 47.1% planned to receive COVID-19 vaccination themselves, 20.6% were unsure, 9.1% reported not planning to receive it and 3.0% declined to answer. Most parents (82.4%) who had been or planned to be vaccinated themselves reported plans to vaccine their youngest child, whereas among parent who were unsure about getting vaccinated or said they did not plan to get vaccinated themselves, only 25.4% and 4.5%, respectively, planned to vaccinate their youngest child (p<0.0001) (Figure 1).

**Figure 1.**
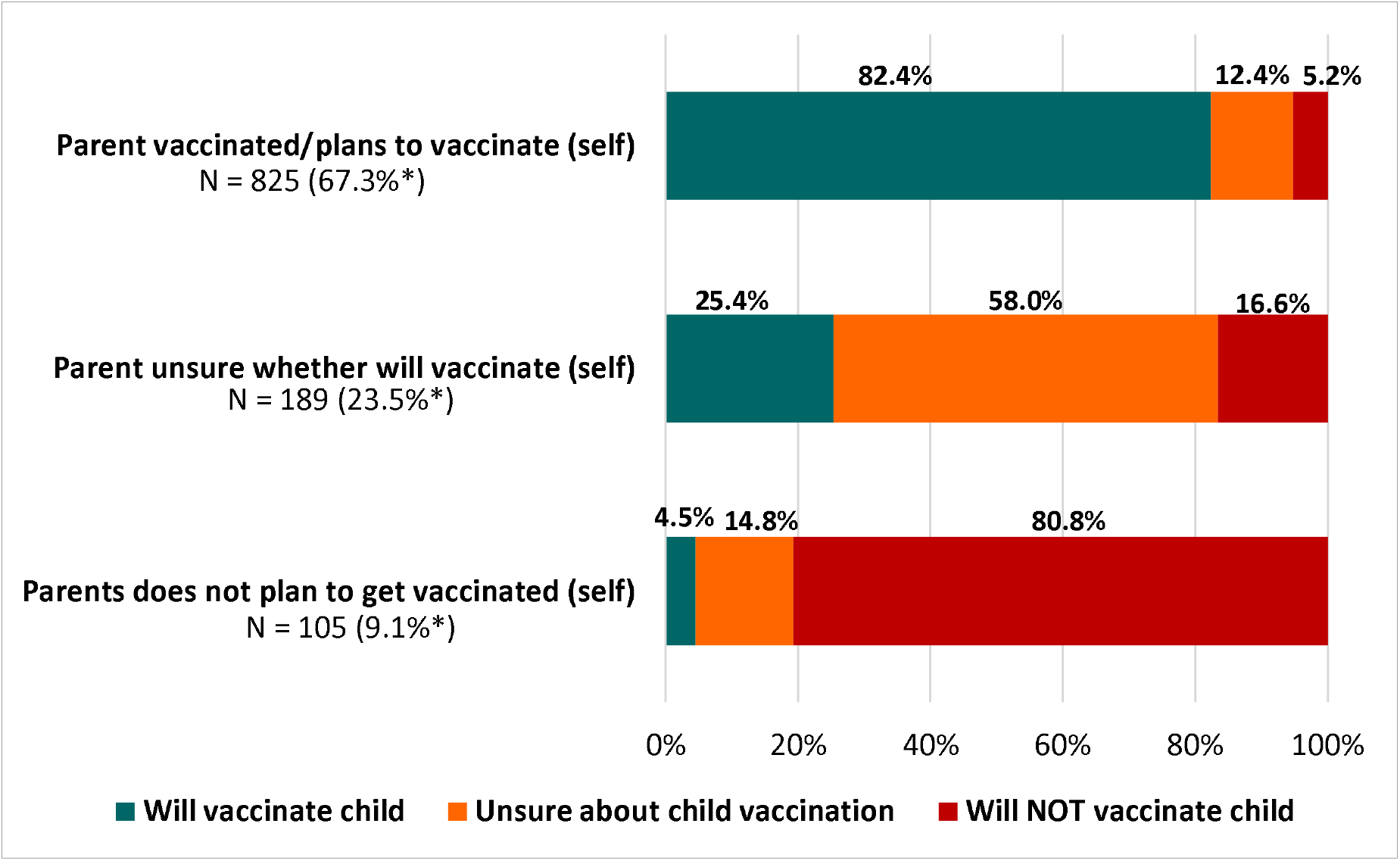
**Parental intentions to vaccinate children in NYC against COVID-19 according to parents’ own vaccination status –** March 9-April 8, 2021 * Survey weights applied to estimate proportions of NYC parents

## DISCUSSION

In March 2021, approximately 60% of parents in NYC reported plans to vaccinate children ≤12 years against COVID-19 when vaccines are approved for pediatric populations. Among parents expressing vaccine hesitancy, the primary concerns were safety and effectiveness, particularly for non-Hispanic Black and Hispanic parents, as well as perceived lack of need. Female and non-Hispanic Black parents were least likely to report intending to vaccinate their children, and we observed a strong correlation between parental willingness to get vaccinated and reported intentions to vaccinate children. Among NYC parents, 67% reported they had been vaccinated or planned to get vaccinated themselves.

Willingness of parents to vaccinate children when a COVID-19 vaccine is approved for pediatric populations will be crucial for protecting pediatric populations, their communities and households from infection. While trials in younger children have not yet been completed, studies in young adolescents (12-15 years) have shown up to 100% efficacy suggesting that the vaccine will be highly effective.^14^ High uptake of efficacious vaccines in pediatric populations may also contribute to lower infections in adults and will help stop community spread, as has been observed with other illnesses.^2^ Decreasing risk of infections in children and curtailing transmission of COVID-19, will also have the added benefit of allowing children to return to normal activities, including school and other services and programs, which have been limited during the epidemic causing significant harm to children and parents.^15^

Our findings indicate that while a majority of parents intend to vaccinate their children for COVID-19, roughly 40% will not or are unsure. In order to achieve herd immunity and stop the spread of COVID-19, it is estimated that up to 70% of the US population may need to be vaccinated, including children.^16^ If vaccine acceptability does not improve, herd immunity may not be reached and COVID-19 may continue circulating among the unvaccinated. In our survey, we found that the most commonly held concerns reported by parents for not wanting to vaccinate their child were safety and efficacy, followed by a perceived lack of need for vaccination of children. This information suggests that greater efforts are needed to help parents understand that vaccines are safe and the importance of vaccinating children to protect them from COVID-19.

We also identified that female parents and non-Hispanic Black parents in NYC were more likely to report vaccine hesitancy. In a survey conducted across the US at the same time, we also found that female parents expressed more hesitancy about COVID-19 vaccination for children.^17^ These findings are consistent with surveys asking adults about their own vaccine intentions conducted in 2020, in which women and Black adults in the US were more likely to report vaccine hesitancy compared to other groups.^18,19^ We did not find that lower income and less education were associated with greater vaccine hesitancy among parents in NYC which is somewhat different from our national survey results, and also different from studies of general vaccine hesitancy measured prior to the epidemic.^17,20^ Targeted efforts to understand the concerns of mothers and to non-Hispanic Black parents to understand their concerns and to explain the safety and benefits of vaccination are needed and should be undertaken now as we await pediatric vaccines in order to increase uptake when approved.

Finally, our study was unique in examining the concordance between parents’ vaccine intentions for themselves and for their child. We found high agreement showing that parents who do not want to get vaccinated themselves will also likely not vaccinate their children, and that parents who are unsure about getting vaccinated are also unsure about the vaccine for their children. We do not know from our data whether these findings indicate that shifts in parents’ willingness to get vaccinated themselves will lead to acceptance of vaccination for children. Further studies are needed to better understand this finding.

Our study was limited to parents in NYC and to children ≤12 years and may not be generalizable to other areas or adolescents. Data were self-reported and subject to response bias, and our survey was conducted online, excluding parents without internet access.

## Conclusions

Our study provides important information about parental acceptability of the COVID-19 vaccine for children in the most densely populated and one of the most racially and ethnically diverse cities in the US. Our findings also suggest that pediatric vaccine hesitancy is strongly tied to parental vaccine hesitancy. Targeted interventions are needed to increase uptake of COVID-19 vaccines for children, including educating parents on the safety and importance of COVID-19 vaccination for children.

## Data Availability

The survey data will be available upon request

